# Patterns of Hepatocellular Carcinoma in Patients with or without Liver Cirrhosis using Computed Tomography in Tanzania: A hospital-based cross-sectional study

**DOI:** 10.1101/2025.05.15.25327588

**Authors:** Stephen Z. Gondwe, Irene H. Mhalu, Jay Lodhia, Twalib A. Ngoma

**Affiliations:** Department of Radiology and Imaging, Muhimbili University of Health and Allied Sciences, P O Box 65001, Dar es Salaam, Tanzania; Department of Radiology, Kilimanjaro Christian Medical Centre, P O Box 3010, Moshi, Tanzania; Department of Radiology and Imaging, Muhimbili National Hospital, P O Box 65000, Dar es Salaam, Tanzania; Department of General surgery, Kilimanjaro Christian Medical Centre, P O Box 3010, Moshi, Tanzania; Faculty of Medicine, Kilimanjaro Christian Medical University College, P O Box 2240, Moshi, Tanzania

**Keywords:** Hepatocellular Carcinoma, Liver Cirrhosis, Hepatitis B, Computed Tomography, Tanzania

## Abstract

**Background:** Hepatocellular Carcinoma (HCC) is the most common primary liver cancer typically occurring in the background of chronic liver disease and cirrhosis. Atypical occurrence of HCC is also present in non-cirrhotic livers.

**Objective:** The aim of this study was to determine the patterns of Hepatocellular carcinoma in patients with or without liver cirrhosis using Computed Tomography (CT) at Muhimbili National Hospital

**Methodology:** A retrospective cross-sectional study was done comparing the CT patterns of clinically suspected cases of HCC who underwent CT imaging at Muhimbili National Hospital from April 2021 to February 2022.

**Results:** A total of 73 cases were enrolled comprising of 78.1% (n = 57) HCC patients with Liver cirrhosis (HCC-LC) and 21.9% (n = 16) HCC patients without Liver cirrhosis (HCC-WLC). HCC-LC presented at a median age of 40 while HCC-WLC presented at a median age of 38. The male-to-female ratio of HCC-LC was 4:1 while that of HCC-WLC was 3:1. Abdominal pain located in the right upper quadrant was the major clinical symptom in both HCC-LC and HCC-WLC (n = 68, 93.2%). The most frequent identified risk factor in both HCC-LC and HCC-WLC was Hepatitis B (n = 40, 54.8%). On CT imaging, multifocal lesions were associated with HCC-LC while solitary lesions were associated with HCC-WLC (P = 0.01). Ill-defined lesions were associated with HCC-LC while well-defined lesions were associated with HCC-WLC (P < 0.01). Lesions in HCC-LC were also more likely to occur in Segment 2, 3 and 4 (P = 0.002, P = 0.003, P = 0.025 respectively).

**Conclusion:** Hepatocellular Carcinoma presents at an earlier age in Tanzania as compared to North America, Europe and Asia. On CT imaging, HCC in patients with liver cirrhosis appears as multifocal and ill-defined lesions favoring the left liver lobe while HCC in patients without liver cirrhosis appears as solitary and well-defined lesions.

## INTRODUCTION

Hepatocellular carcinoma (HCC) is the most common primary liver cancer and a leading cause of death among patients with liver cirrhosis. It typically occurs in a cirrhotic liver but in 20% of the time, it may occur in a non-cirrhotic liver. HCC has a rapid progression by local extension and metastases with a low life expectancy of between 6 to 20 months (1–3).

According to WHO, Liver cancer is the sixth common cancer and third leading cause of cancer-related deaths globally. In Tanzania, liver cancer is among the three leading causes of cancer-related deaths affecting 11.3% of males and 5.1% of females dying from cancer (4–6).

The pathogenesis of HCC is a complex multistep process that involves a variety of pathways which can be modified by environmental and genetic-related factors leading to the transformation of normal Hepatocytes into malignant cells (3,7,8).

The main risk factors associated with HCC are Hepatitis B (HBV) and Hepatitis C (HCV). The prevalence of HBV and HCV in Tanzania is estimated to be around 6-9% and 2-3% respectively. Other risk factors associated with HCC include alcohol abuse, Schistosomiasis, Aflatoxins and Tobacco use (7,9–13).

The clinical presentation of HCC includes right upper abdominal pain, right hypochondrial swelling, jaundice, weight loss, fatigue, anemia, ascites and hematemesis. If the cancer is advanced, patients may present with metastasis to the lungs, abdominal lymph nodes, bone, adrenal glands and peritoneum (14– 17).

Clinical criteria for HCC diagnosis consists of presence of background liver disease, elevated tumor markers and suggestive imaging findings. On triphasic CT imaging, HCC classically exhibits early arterial phase enhancement and washout in the portal venous and delayed phases. Biopsy is the gold standard for diagnosing HCC although it is rarely possible in Tanzania due to limited areas with interventional procedures (18–21).

Alfa fetoprotein (AFP) is elevated in 75% of cases. AFP is a serum glycoprotein that is produced by the fetal liver and embryonic yolk sac during pregnancy and declines to less than 10ng/ml after birth. AFP of greater than 200-400 ng/ml in adults is often considered diagnostic of HCC in the context of suggestive imaging findings (22–26).

Management of HCC involves tumor resection in cases without liver cirrhosis and liver transplantation in cases with liver cirrhosis. Alternative treatment includes trans-arterial Chemoembolization and Radiofrequency ablation for patients not fit for resection nor transplantation (27).

In Tanzania, HCC commonly presents at an advanced stage and in younger patients as compared to Europe and USA. This can be prevented by awareness and reduced exposure to predisposing risk factors. The objectives of this study were to establish the radiological patterns and characteristics of patients with HCC (5,11).

## METHODOLOGY

### Study design

This was a retrospective hospital based cross sectional study of patients clinically suspected with Hepatocellular carcinoma who underwent triphasic CT imaging at the Radiology Department of Muhimbili National Hospital from April 2021 to February 2022.

### Study setting

Muhimbili National Hospital is the largest referral hospital in Tanzania, located in the Ilala municipality of Dar es Salaam. It serves as a treatment facility, research center and university teaching hospital. The Radiology Department is equipped with Ultrasound, X-Ray, CT and MRI machines.

### Study population

The study included all patients clinically suspected with Hepatocellular Carcinoma who underwent triphasic CT imaging during the study period and fulfilled the inclusion criteria.

### Inclusion criteria

□ All patients with clinically suspected Hepatocellular Carcinoma who underwent triphasic CT imaging at Muhimbili National Hospital and satisfied clinical criteria for HCC
□ All patients with clinically suspected Hepatocellular Carcinoma who underwent triphasic CT imaging at Muhimbili National Hospital and had confirmatory Biopsy

### Exclusion criteria

□ Patients whose clinical data was incomplete and/or inconclusive

### Sampling Technique

Convenient non-probability sampling was used whereby all patients with features of HCC on CT imaging during the study period and who met inclusion criteria were included in the study. HCC diagnosis was confirmed based on standard published criteria (22). Histological sampling was unfortunately only done in conditions when non-invasive parameters were not diagnostic. Treatment outcomes were not included as MNH has limited treatment options for advanced HCC.

### Sample size estimation

By using the sample size estimation formula, n = Z^2^P(1-P)/e^2^ where; n is the minimum sample size, Z is the standard deviation corresponding to 95% confidence interval (z=1.96), e is the desired precision (set at 0.05), P is 4.6% (proportion of HCC at BMC)(15)

Therefore; n = ((1.96)^2^ x 0.046) (1 – 0.046) / (0.05)^2^ = 68

### Dependent variables

Parenchymal nodularity, Caudate Lobe enlargement, Fatty change, Number of Lesions, Tumor margins, Consistency, Ascites, Lymphadenopathy, Metastases, Portal vein and Inferior vena cava thrombosis

### Independent variables

Age, Sex, Hepatitis B, Hepatitis C, HIV and history of Schistosomiasis

### Data collection

A pilot study was first done at MNH and thereafter a structured data collection tool was made. Triphasic CT images were acquired by a 128 slice dual source CT scanner using bolus tracking method. Clinical history and Laboratory findings were obtained from hospital files and patient database. CT images were interpreted by the primary investigator and reviewed by two experienced radiologists.

### Data analysis

Analysis was done using the Statistical Package for Social Sciences (SPSS) version 28. Median and range were calculated for continuous variables whereas proportions were computed for categorical variables. The chi-square (x^2^) test was used to determine association between predictor and outcome where P < 0.05 was considered as level of significance. Incomplete data was listed as unknown or unspecified.

### Ethical consideration

A formal ethical approval was obtained from the Institutional Review Board (IRB) of the Muhimbili University of Health and Allied Sciences. Permission to conduct the study at MNH in Radiology department was obtained from necessary authorities. Patient’s information was kept confidential and names were not used.

## RESULTS

A total of 73 HCC cases were obtained in the MNH radiology department between April 2021 and February 2022 of which 71.9% (n = 57) were HCC patients with liver cirrhosis (HCC-LC) and 28.07% (n = 16) were HCC patients without liver cirrhosis (HCC-WLC).

The study patients had an age distribution ranging from 22 to 71 years in HCC-LC and from 18 to 84 years in HCC-WLC. The most affected age group in both HCC-LC and HCC-WLC was of 31-40 years (38.4%, n = 28). Males were the most affected sex in both HCC-LC and HCC-WLC with male-to-female ratios of 1:4 in HCC-LC and 1:3 in HCC-WLC. Right upper abdominal pain and abdominal swelling were associated with HCC-LC but were generally also the most frequently identified symptoms (Table 1).

**Table 1:**
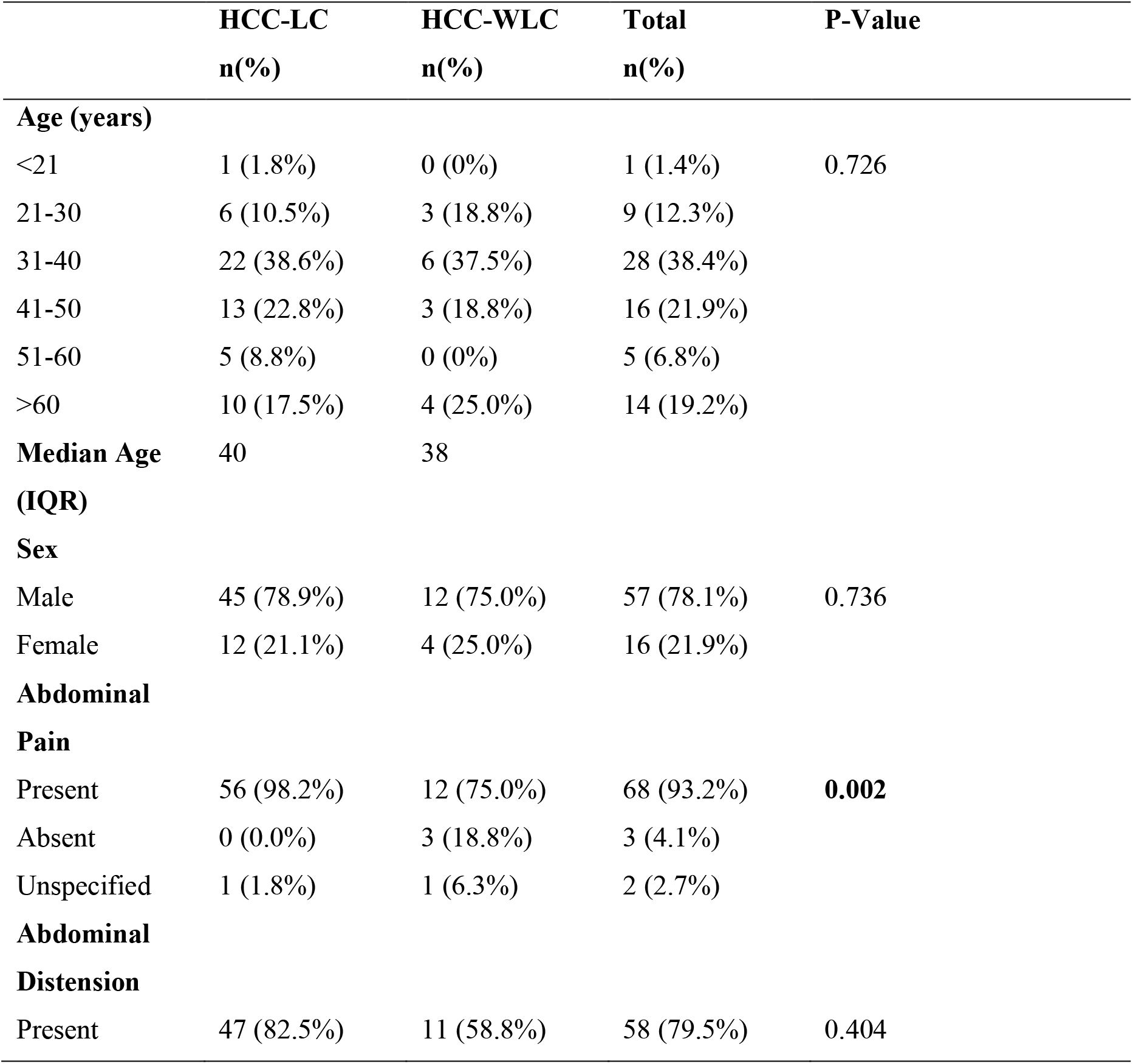
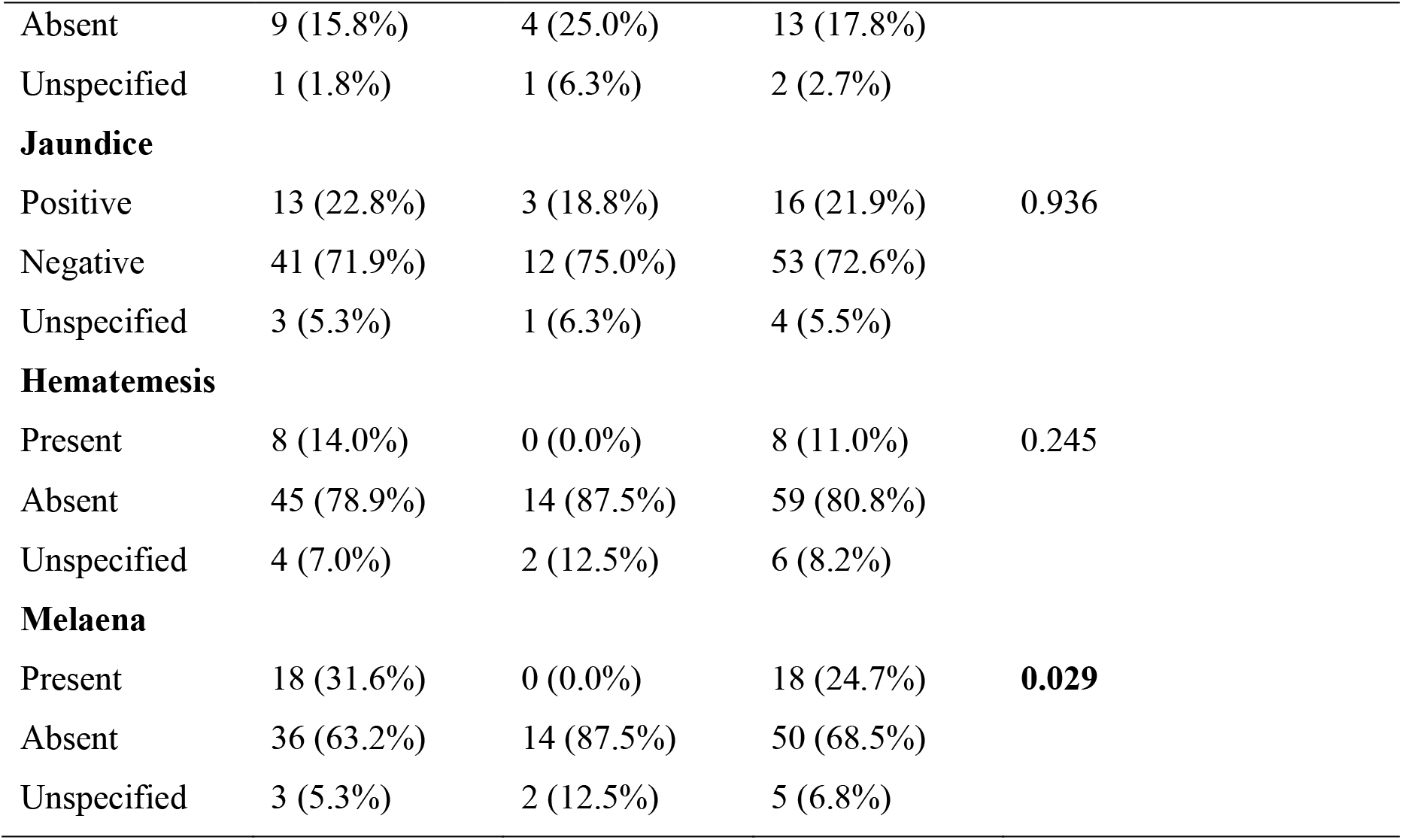
Socio-demographic and Clinical Characteristics of the study participants (N=73)

The most frequently identified predisposing risk factor was Hepatitis B (54.8%, n=40). Hepatitis C was not common despite being investigated in only 52 study patients. History of schistosomiasis was common in patients with history of working in rice plantations. Other risk factors identified were alcohol and smoking but quantification was difficult because of inconsistent documentation (Table 2).

**Table 2:**
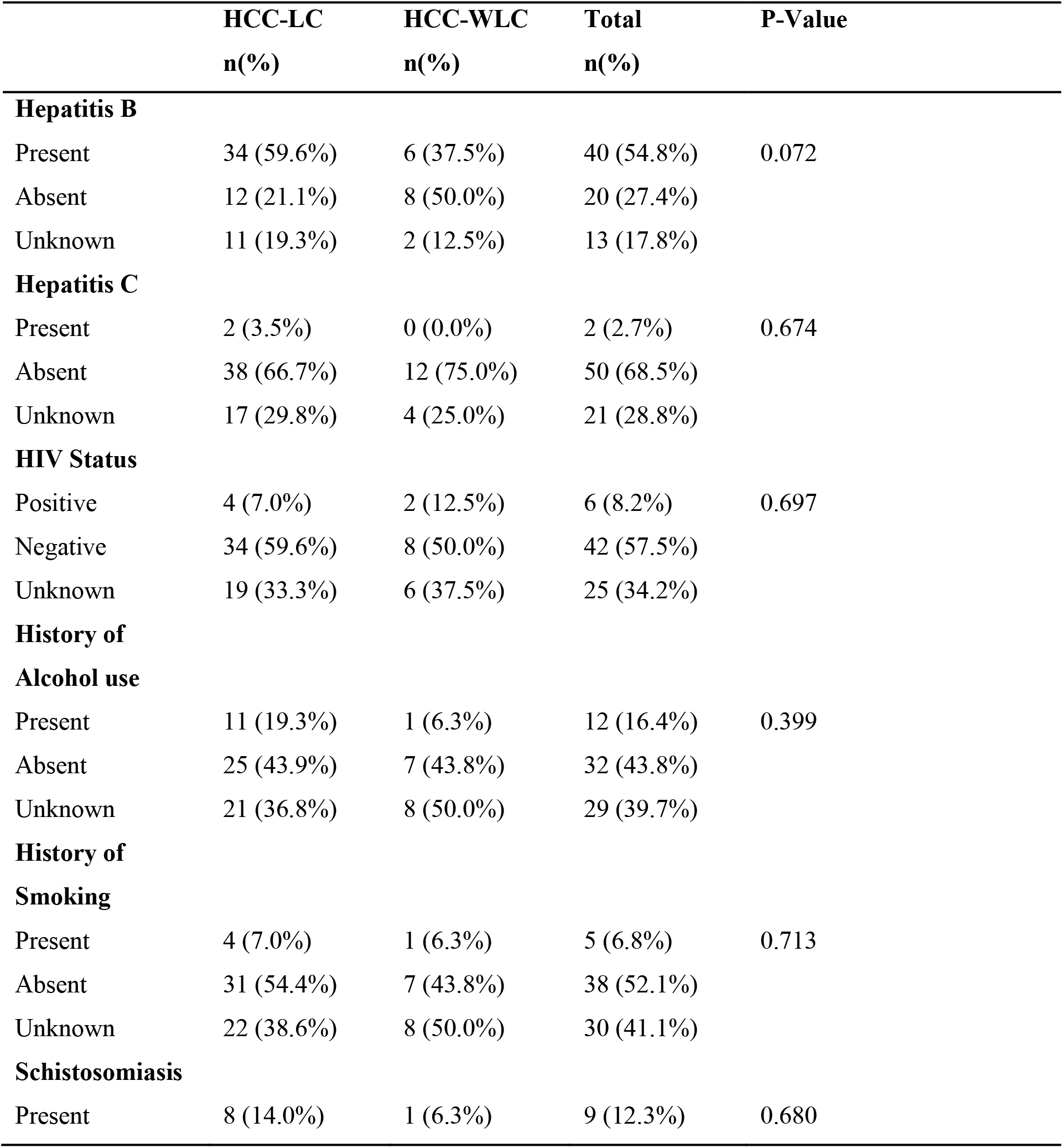
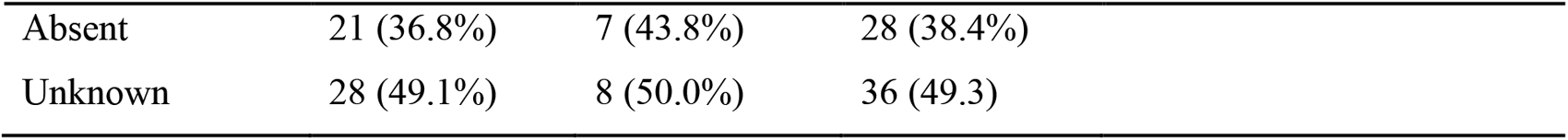
Predisposing risk factors in the study participants (N=73)

|  | HCC-LC | HCC-WLC | Total | P-Value |
| --- | --- | --- | --- | --- |
|  | n(%) | n(%) | n(%) |  |
| **Hepatitis B** |  |  |  |  |
| Present | 34 (59.6%) | 6 (37.5%) | 40 (54.8%) | 0.072 |
| Absent | 12 (21.1%) | 8 (50.0%) | 20 (27.4%) |  |
| Unknown | 11 (19.3%) | 2 (12.5%) | 13 (17.8%) |  |
| **Hepatitis C** |  |  |  |  |
| Present | 2 (3.5%) | 0 (0.0%) | 2 (2.7%) | 0.674 |
| Absent | 38 (66.7%) | 12 (75.0%) | 50 (68.5%) |  |
| Unknown | 17 (29.8%) | 4 (25.0%) | 21 (28.8%) |  |
| **HIV Status** |  |  |  |  |
| Positive | 4 (7.0%) | 2 (12.5%) | 6 (8.2%) | 0.697 |
| Negative | 34 (59.6%) | 8 (50.0%) | 42 (57.5%) |  |
| Unknown | 19 (33.3%) | 6 (37.5%) | 25 (34.2%) |  |
| **History of Alcohol use** |  |  |  |  |
| Present | 11 (19.3%) | 1 (6.3%) | 12 (16.4%) | 0.399 |
| Absent | 25 (43.9%) | 7 (43.8%) | 32 (43.8%) |  |
| Unknown | 21 (36.8%) | 8 (50.0%) | 29 (39.7%) |  |
| **History of Smoking** |  |  |  |  |
| Present | 4 (7.0%) | 1 (6.3%) | 5 (6.8%) | 0.713 |
| Absent | 31 (54.4%) | 7 (43.8%) | 38 (52.1%) |  |
| Unknown | 22 (38.6%) | 8 (50.0%) | 30 (41.1%) |  |
| **Schistosomiasis** |  |  |  |  |
| Present | 8 (14.0%) | 1 (6.3%) | 9 (12.3%) | 0.680 |

|  |  |  |  |
| --- | --- | --- | --- |
| Absent | 21 (36.8%) | 7 (43.8%) | 28 (38.4%) |
| Unknown | 28 (49.1%) | 8 (50.0%) | 36 (49.3) |

On CT imaging, multifocal lesions were associated with HCC-LC while solitary lesions were associated with HCC-WLC (P = 0.01). Ill-defined lesions were associated with HCC-LC while well-defined lesions were associated with HCC-WLC (P < 0.01). Additionally, Ascites (P-value = 0.001) and Portal Venous Thrombosis (P-value = 0.001) were associated with HCC-LC. Criteria used to determine metastatic lymph nodes was short axis diameter greater than 1 cm (Table 3).

**Table 3:** CT image findings in the study participants (N=73)

|  | HCC-LC | HCC-WLC | Total | P-Value |
| --- | --- | --- | --- | --- |
|  | n(%) | n(%) | n(%) |  |
| Number of lesions |  |  |  |  |
| Single | 13 (22.8%) | 9 (56.3%) | 22 (30.1%) | 0.010 |
| Multiple | 44 (77.2%) | 7 (43.8%) | 51 (69.9%) |  |
| Tumor margins |  |  |  |  |
| Well-defined | 25 (43.9%) | 15 (93.8%) | 40 (54.8) | 0.001 |
| Ill-defined | 32 (56.1%) | 1 (6.3%) | 33 (45.2%) |  |
| Consistency |  |  |  |  |
| Solid | 35 (61.4%) | 7 (43.8%) | 42 (57.5%) | 0.207 |
| Necrotic | 22 (38.6%) | 9 (56.3%) | 31 (42.5%) |  |
| Ascites |  |  |  |  |
| Present | 43 (75.4%) | 5 (31.3%) | 48 (65.8%) | 0.001 |
| Absent | 14 (24.6%) | 11 (68.8%) | 25 (34.2%) |  |
| Portal Vein Thrombosis |  |  |  |  |
| Present | 27 (47.4%) | 0 (0.0%) | 27 (37.0%) | 0.001 |
| Absent | 30 (52.6%) | 16 (100%) | 46 (63.0%) |  |
| IVC Thrombosis |  |  |  |  |
| Present | 5 (8.8%) | 1 (6.3%) | 6 (8.2%) | 0.746 |
| Absent | 52 (91.2%) | 15 (93.8%) | 67 (91.8%) |  |
| Abdominal Lymphadenopathy |  |  |  |  |
| Enlarged | 15 (26.3%) | 4 (25.0%) | 19 (26.0%) | 0.916 |
| Not enlarged | 42 (73.7%) | 12 (75.0%) | 54 (74.0%) |  |

Tumors arising from liver segment 2, 3 and 4 showed an association with HCC-LC suggesting that HCC in presence of liver cirrhosis was most likely to arise from the left liver lobe (Table 4).

**Table 4:** Liver Segments involved by HCC (N=73)

|  | HCC-LC<br>n(%) | HCC-WLC<br>n(%) | Total<br>n(%) | P-Value |
| --- | --- | --- | --- | --- |
| Segment 1 |  |  |  |  |
| Involved | 30 (52.6%) | 6(37.5%) | 36 (49.3%) | 0.285 |
| Not involved | 27 (47.4%) | 10 (62.5%) | 37 (50.7%) |  |
| Segment 2 |  |  |  |  |
| Involved | 39 (68.4%) | 4 (25.0%) | 43 (58.9%) | 0.002 |
| Not involved | 18 (31.6%) | 12 (75.0%) | 30 (41.1%) |  |
| Segment 3 |  |  |  |  |
| Involved | 38 (66.7%) | 4 (25.0%) | 42 (57.5%) | 0.003 |
| Not involved | 19 (33.3%) | 12 (75.0%) | 31 (42.5%) |  |
| Segment 4 |  |  |  |  |
| Involved | 39 (68.4%) | 6 (37.5%) | 45 (61.6%) | 0.025 |
| Not involved | 18 (31.6%) | 10 (62.5%) | 28 (38.4%) |  |
| Segment 5 |  |  |  |  |
| Involved | 45 (78.9%) | 13 (81.3%) | 58 (79.5%) | 0.840 |
| Not involved | 12 (21.1%) | 3 (18.8%) | 15 (20.5%) |  |
| Segment 6 |  |  |  |  |
| Involved | 45 (78.9%) | 11 (68.8%) | 56 (76.7%) | 0.394 |
| Not involved | 12 (21.1%) | 5 (31.3%) | 17 (23.3%) |  |
| Segment 7 |  |  |  |  |
| Involved | 46 (80.7%) | 10 (62.5%) | 56 (76.7%) | 0.128 |
| Not involved | 11 (19.3%) | 6 (37.5%) | 17 (23.3%) |  |
| Segment 8 |  |  |  |  |
| Involved | 46 (80.7%) | 12 (75.0%) | 58 (79.5%) | 0.618 |
| Not involved | 11 (19.3%) | 4 (25.0%) | 15 (20.5%) |  |

Extrahepatic metastases were found in 38% (n = 28) of the study sample. The identified sites of metastases were the Lungs, Bones and Adrenal glands. Lungs were the most common extrahepatic site of metastasis (Figure 2).

**Figure 2:**
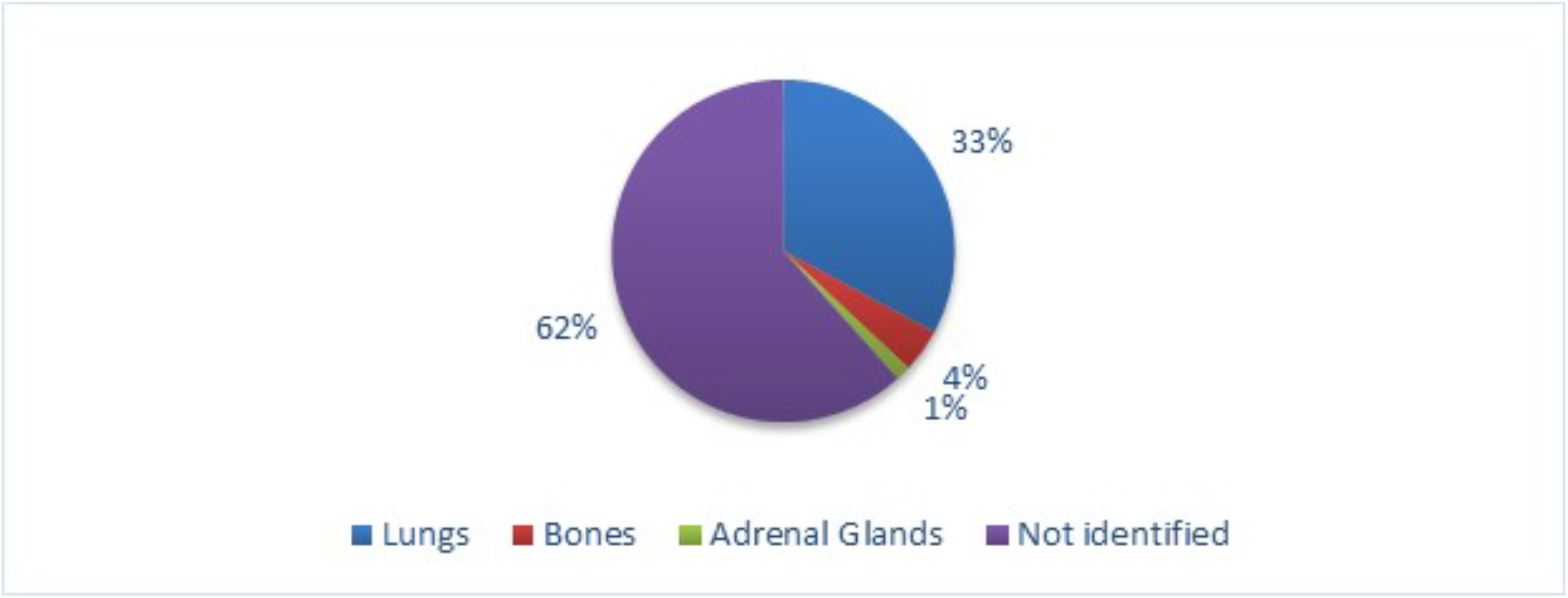
Pie chart showing frequency of sites involved in Metastatic HCC (N=73)

## DISCUSSION

A common modal age group of 31-40 years was observed for HCC in MNH. This is lower than the modal age group of 41-50 years reported previously for HCC at BMC. This discrepancy may be attributed to differences in catchment areas or sample size. In general, HCC has an earlier onset in Tanzania as compared to Western and Asian countries (15,28–31).

Males were the most affected gender with a male-to-female ratio of 3:1. This ratio is in agreement with previous studies and is as an indicator of the higher predisposition of males to HCC. The male-to-female ratio for HCC patients with and without liver cirrhosis were 4:1 and 3:1 respectively. This is similar to previous studies that report a reduced male-to-female ratio in HCC patients without liver cirrhosis (28,32– 34).

The higher incidence of HCC in males is attributed to a mediatory effect of Testosterone in males and protective effect of Estrogen in females. Testosterone mediates the inhibition of Adiponectin secretion which leads to unchecked hepatic cancer cell proliferation while Estrogen inhibits production of interleukin-6 from Kupffer cells exposed to necrotic hepatocytes thus preventing induction of hepatocarcinogenesis (35,36).

The common clinical symptoms in both HCC-LC and HCC-WLC were right upper abdominal pain and abdominal distension. Right upper abdominal pain and Melaena were also more common in HCC-LC than in HCC-WLC. This is partly similar to the study done at BMC where right upper abdominal pain was the most identified symptom (15).

Hepatitis B was the most identified risk factor in both HCC patients with and without liver cirrhosis. This is in agreement with multiple studies portraying Hepatitis B as the predominant risk factor in Africa. The usual pathway from Hepatitis B infection to HCC is usually through cirrhosis but Hepatitis B can also cause HCC prior to cirrhosis through PIK3CA gene mutations which lead to sporadic cancer (33,37)

On CT imaging, HCC with liver cirrhosis presented as multifocal and ill-defined lesions assuming an infiltrative pattern while HCC without liver cirrhosis presented as solitary and well-defined lesions. This is similar to a study done in India and may be due to changes in architecture during cirrhosis which facilitates an infiltrative-like spread of cancer cells. (30,31,33).

There was an association between lesions in segment 2, 3 and 4 of the left liver lobe and the presence of liver cirrhosis whereas in absence of liver cirrhosis, lesions were frequently found in segment 5, 6, 7 and 8 of the right liver lobe. The predominance of lesions in the right lobe in HCC-WLC is similar to findings of a study done in India where 83% of cases without liver cirrhosis had lesions only in the right lobe (31).

Ascites was a dominant feature in HCC patients with liver cirrhosis. Despite association being established, it is important to note that ascites may be caused by both tumoral and cirrhotic factors. A study in Taiwan found that ascites was associated with a larger tumor burden which in this study may be supported by the larger proportion of HCC-LC cases with a liver surface tumor involvement of >50% (40).

A higher incidence of Portal Vein Thrombosis was noted in HCC patients with liver cirrhosis. Reviewed literature proposes that Portal Vein Thrombosis has a propensity of occurring in patients with infiltrative or massive type HCC. Blood hypercoagulability and other flow alterations that occur during the process leading up to Cirrhosis are likely factors that contribute to this finding (38,41).

The identified sites of metastases, in order of occurrence, were the lungs, bones and adrenal glands. This order is in agreement with the metastatic patterns found in Germany and Italy but in contrast to these studies, there was no association to suggest higher metastatic occurrence in HCC patients without liver cirrhosis as compared to those with liver cirrhosis (17,29,42).

## LIMITATIONS

This was a retrospective study and was prone to encountering incomplete or inaccurate medical records. The study being hospital-based could lead to selection bias and may not represent the entire population. Variability in diagnostic criteria and imaging techniques could also affect the consistency of the findings.

## CONCLUSION

In conclusion, this hospital based study reveals distinct patterns of hepatocellular carcinoma in patients with and without liver cirrhosis. Patients with liver cirrhosis exhibit a higher prevalence of multifocal tumors and portal vein involvement resulting in a poorer prognosis compared to those without cirrhosis. These results highlight the importance of early detection and tailored treatment strategies to improve the outcome of HCC in Tanzania.

## Data Availability

All data produced in the present study are available upon reasonable request to the authors

## ABBREVIATIONS

AFP: Alpha Fetoprotein
BMC: Bugando Medical Centre
CT: Computed Tomography
HBV: Hepatitis B Virus
HCC: Hepatocellular Carcinoma
HCC-LC: Hepatocellular Carcinoma with Liver Cirrhosis
HCC-WLC: Hepatocellular Carcinoma without Liver Cirrhosis
HCV: Hepatitis C Virus
HIV: Human Immunodeficiency Virus
IQR: Interquartile Range
MNH: Muhimbili National Hospital
MRI: Magnetic Resonance Imaging
WHO: World Health Organization

## Conflict Of Interest

The authors declare no conflict of interest.

## Authors’ Contributions

GSZ designed the study, collected data, entered data in SPSS for analysis and interpreted the results, wrote the report and drafted this manuscript. MIH, JL and NTA participated in revising the report and editing the draft manuscript prior to submission. All authors have proofread and approved the final manuscript.

## Acknowledgement

I would like to thank God for giving me the strength to complete this work. I would also like to express my gratitude to the departments of radiology at MUHAS and MNH for mentoring me during my coursework and research.

## Notes

### Competing Interest Statement

The authors have declared no competing interest.

### Funding Statement

This study did not receive any funding

### Author Declarations

Institutional Review Board of Muhimbili University of Health and Allied Sciences and Muhimbili National Hospital gave ethical approval for this work

